# Causal Inference and COVID-19 Nursing Home Patients: Identifying Factors That Reduced Mortality Risk

**DOI:** 10.1101/2021.11.18.21266489

**Authors:** Amina Ahmed, Robert Goldberg, Joseph Swiader, Zachary A.P. Wintrob, Margaret Yilmaz

**Affiliations:** Prosper Digital Therapeutics, Inc; Care One; Prosper Digital Therapeutics; Roaketen, Inc

## Abstract

Less than 1% of the US population lives in long-term care facilities, yet this subset of the population accounts for 22% of total COVID-19 related deaths.^1^ Because of a lack of experimental evidence to treat COVID-19, analysis of real-world data to identify causal relationships between treatments/policies to mortality and morbidity among high-risk individuals is critical. We applied causal inference (CI) analysis to longitudinal patient-level health data of 4,091 long-term care high-risk patients with COVID-19 to determine if any actions or therapies delivered from January to August of 2020 reduced COVID-19 patient mortality rates during this period.

Causal inference findings determined that certain supportive care interventions caused reduced mortality rates for nursing home residents regardless of severity of disease (as measured by oxygen saturation level, presence of pneumonia and organ failure), comorbidities or social determinants of health such as race, age, and weight.^2^ While we do not address the biological mechanisms associated with specific medical interventions and their impact on mortality, this analysis suggests methods to validate and optimize treatment protocols using domain knowledge and causal inference analysis of real-world data across patient populations and care settings.

## Introduction

Throughout the COVID-19 pandemic, nursing home residents have died of COVID-19 at substantially higher rates than people in hospitals or living at home. As the pandemic began in early 2020, the death rate for nursing home patients infected with the SARS-CoV2 virus was over 30 percent. By August 30, 2020, the COVID-19 death rate in nursing homes had dropped to 18.2 percent, and the number of new COVID-19 cases declined as well.^3^ COVID-19 mortality rates declined during the first wave of the pandemic (from January through October) among nursing home residents and the general population. As of September 2020, “people in long-term care facilities were 8 percent of coronavirus cases, but 45 percent of all COVID-19 death.”^4^

There is a limited amount of COVID-19 research seeking to identify treatments or detection steps associated with reduced mortality. While randomized controlled clinical trials can be used to establish whether a treatment causes a reduction in mortality, such studies are often difficult to conduct or may be unethical. A recent study of mortality rates among nursing home residents concluded that mortality risk “declined for residents with both symptomatic and asymptomatic infections and for residents with both high and low clinical complexity.”^5^ The authors surmised that a change in the transmissibility and severity of the virus may have caused this shift and did not find any interventions that were associated with reduced mortality.^6^ Other studies have attempted to identify specific treatment regimens or detect strategies that might have conveyed such a benefit. A Cochrane Collaboration meta-analysis found that “based on currently available data, neither absence nor presence of signs or symptoms are accurate enough to rule in or rule out COVID-19. The presence of anosmia or ageusia may be useful as a red flag for COVID-19. The presence of fever or cough, given their high sensitivities, may also be useful to identify people for further testing. Prospective studies in an unselected population presenting to primary care or hospital outpatient settings, examining combinations of signs and symptoms to evaluate the syndromic presentation of COVID-19, are still urgently needed.”

In the absence of randomized controlled trials, the analysis of observational longitudinal data should be considered for evaluating the impact of specific treatments and interventions on COVID-19 mortality. However, as stated by Judea Pearl PhD, pioneer of Bayesian statistics and causal inference, “Interventional questions cannot be answered from purely observational information (i.e., from statistical data alone).”^7^ A recent article on causal inference and clinical decision-making observes, “Prediction calculates a future event in the absence of any action or change; intervention presumes an enacted choice that may influence the future, which requires consideration of the underlying causal structure.”^8^ Predictions alone do not provide clinicians with the ability to determine whether certain interventions or policies have improved outcomes. This analysis sought to identify the *causal* effects of actions taken to treat high-risk COVID-19 patient with regard to the reduction of mortality risk. Specifically, we asked if various treatments of COVID-19 patients caused a lower rate of mortality.

## Methods

### Data Source and Population

The source data for the analysis was provided by CareOne and consisted of clinical records for 4,091 COVID-19 patients diagnosed and treated from January 2020 through August 2020. Data were de-identified and determined by an independent institutional review board as nonhuman subject research. The cohort was composed of both inpatient and outpatient residents at 55 nursing and assisted living facilities in the northeastern United States. All continuous variables were standardized prior to model fitting.

We analyzed data with several hundred variables for every patient including demographics (age, gender, race, region, insurance status), preexisting conditions such as diabetes, obesity, BMI, heart disease, cancer, as well up to twenty-six different lab results, oxygen, incentive spirometry, etc. Patient data also included vital signs, medications and whether the patient was being provided hospice or palliative care or were in an assisted living, post-acute care, or long-term care setting.

Of the 4,091 residents with confirmed COVID-19 infections in the study population, 2,277 (55.7%) were female. Most COVID-19 positive cases identified as Caucasian, 2,630 (64.3%) with 375 (9.2%) identifying as African American. Of the total COVID-19 patients evaluated during the study time frame there 1,204 new admissions and re-admissions that tested positive for COVID-19 after being admitted from hospitals. All COVID-19 infections in this subset occurred within 14 days of admission. Baseline characteristics of the study population and survivors subgroups are shown in Tables 1 and 2, respectively. Table 3 shows the distribution of comorbidities among the study population.

**Table 1:** Select COVID-19 Patient Demographics.

| Category | Mean | Min | Max | State | N | % |
| --- | --- | --- | --- | --- | --- | --- |
| Age | 79 | 23 | 109 | New Jersey | 2,188 | 53.5% |
| BMI | 26.6 | 11.3 | 83.1 | Massachusetts | 1,471 | 36.0% |
| Height | 67.5 | 45.0 | - | New York | 120 | 2.9% |
| IBW | 133.4 | 65.2 | 214.0 | Maryland | 54 | 1.3% |
|  |  |  |  | Pennsylvania | 20 | 0.5% |
| **Category** |  | **N** | **%** | Connecticut | 15 | 0.4% |
| Females |  | 2,275 | 55.6% | Rhode Island | 13 | 0.3% |
| Males |  | 1,816 | 44.4% | New Hampshire | 12 | 0.3% |
|  |  |  |  | Florida | 11 | 0.3% |
| **Race** |  | **N** | **%** | Virginia | 5 | 0.1% |
| Asian |  | 89 | 2.2% | Maine | 4 | 0.1% |
| Black or African |  | 374 | 9.1% | Vermont | 4 | 0.1% |
| Hispanic or Latino |  | 229 | 5.6% | Delaware | 2 | >0.1% |
| Others |  | 769 | 18.8% | Ohio | 2 | >0.1% |
| White |  | <u>2,630</u> | 64.3% | Texas | 2 | >0.1% |
| Total |  | 4,091 | 100.0% | Washington DC | 1 | >0.1% |
|  |  |  |  | Illinois | 1 | >0.1% |
| **Education** |  | **N** | **%** | Minnesota | 1 | >0.1% |
| Graduate degree |  | 51 | 1.2% | New Mexico | 1 | >0.1% |
| Bachelor's degree |  | 112 | 2.7% | Nevada | 1 | >0.1% |
| Some college |  | 164 | 4.0% | Unknown | <u>163</u> | <u>4.0%</u> |
| High School |  | 511 | 12.5% | Total | 3,928 | 100.0% |
| Less than High |  | 177 | 4.3% |  |  |  |
| Unknown |  | 3076 | 75.2% |  |  |  |
| **Total** |  |  | 100.0% |  |  |  |

**Table 2:** COVID-19 Survivors.

| Category | Sub-category | Value | SD or % |
| --- | --- | --- | --- |
| N = 3,395 |  |  |  |
| Age, mean (sd) |  | 78.2 years | 12.6 years |
| Sex, n (%) | Female | 1,898 | 55.9% |
| Sex, n (%) | Male | 1,497 | 44.1% |
| Race, n (%) | Asian | 69 | 2.0% |
| Race, n (%) | Black of African | 343 | 10.1% |
| Race, n (%) | Hispanic or Latino | 206 | 6.1% |
| Race, n (%) | Others | 621 | 18.3% |
| Race, n (%) | White | 2,156 | 63.5% |
| Temperature to onset* | Days | 2.7 days | 13.7 days |
| COVID-19 | Mild | 100 | 2.9% |
| COVID-19 | Moderate | 72 | 2.1% |
| COVID-19 | Severe | 3,223 | 94.9% |

\*Mean time in days from first 99° F temperature to onset of COVID-19

**Table 3:** COVID-19 Patient Comorbidities.

| Comorbidity | Counts | % Patients with comorbidity |
| --- | --- | --- |
| Hypertension | 1,806 | 44.6% |
| Dementia | 1,362 | 33.3% |
| Behavioral | 1,081 | 26.4% |
| Diabetes | 995 | 24.3% |
| Depression | 943 | 23.1% |
| Obesity | 906 | 22.2% |
| Anemia | 836 | 20.4% |
| Heart disease | 726 | 17.8% |
| Psychosis | 686 | 16.8% |
| Atrial fibrillation | 612 | 15.0% |
| COPD or asthma | 604 | 14.8% |
| Chronic kidney disease | 541 | 13.2% |
| Anxiety | 538 | 13.2% |
| Congestive heart failure | 431 | 10.5% |
| Cerebrovascular | 421 | 10.3% |
| Morbid obesity | 348 | 8.5% |
| Cancer | 346 | 8.5% |
| Peripheral vascular | 346 | 8.5% |
| Kidney failure | 278 | 6.8% |
| Vitamin D deficiency | 175 | 4.3% |
| Parkinson's | 167 | 4.1% |
| Hyponatremia | 135 | 3.3% |
| Hypokalemia | 115 | 2.8% |
| Weight loss > 5 pounds | 63 | 1.6% |
| Other autoimmune | 63 | 1.6% |
| Rheumatoid arthritis | 58 | 1.4% |
| Dehydration | 52 | 1.3% |
| Hyperkalemia | 50 | 1.2% |
| Heart bypass | 41 | 10.0% |
| Lupus | 8 | 0.0% |

### Causal Inference

As Pearl notes, “Graphical models such as Directed Acyclic Graphs (DAGs) can be used for encoding as well as portraying conditional independencies and causal relations.”^9^ Accordingly, we constructed directed acyclic graphs using the patient health record variables to differentiate causal relationships from associations in the data. A DAG clarifies study questions and identifies and illustrates assumptions (in the form of lines or edges) about the relationships between variables (nodes) and reduces bias by separating individual effects and establishing appropriate covariates. The DAG is *directed* in that its lines point from cause to effect (causal effects cannot be bidirectional); and *acyclic* in that no directed path can form a closed loop in a DAG.

DAGs make assumptions and potential causal relationships transparent. Changes to the models are made by holding every other variable constant that might account for the outcome to observe the effect of changing or removing a factor or action.^10^ We generated a DAG for every patient by using lab values as instrumental variables (i.e., variables more strongly associated with the treatment than with the outcome) to eliminate confounding and to identify latent (unobserved) variables to eliminate residual bias carried by unmeasured confounders.^11^

To isolate specific independent effect of medicines or treatment steps, we generated thousands of possible of heterogeneous treatment effects of concurrent pharmacotherapy using Orthogonal Random Forests to control for confounders. For each patient, treatment specific models were trained to predict constant marginal average treatment effects and constant marginals for mortality with a minimum leaf size of 50 and a maximum depth of two.^12^ We then developed and calculated a propensity score for causal relationships for each patient to establish the conditional probability of observed variables. This was accomplished by using treatment specific logistic-regression models to predict constant average treatment effects and constant marginals for specific therapies across the entire patient population. The internal treatment predictions were fit by logistic regression with 3-fold cross-validation. The internal outcome model also implemented 3-fold cross-validation with a ridge classifier with 2 exceptions, the morphine and sennosides outcome models were fit with weighted LASSO.^13^ Analysis was conducted using Python 3.9.0 and the EconML and Scikit-Learn packages.^14^ We measure survival as a Boolean feature representing whether a COVID-19 patient or former COVID-19 patient “died” or “survived”. If a patient died within 120 days of a covid diagnosis the mortality outcome target was “1”, otherwise it was “0”. ^15^

## Results

### Mortality

Table 4 provides morality rates for the entire population (17.3%). The COVID-19 mortality rate of non-hospice COVID-19 patients between January and August 2020 was 9.9% percent. During this period, CareOne took in several hundred medically complex COVID-19 patients from hospitals and other facilities at the request of the New Jersey State Department of Health, including those who were receiving hospice and palliative care in other settings.^16^ By comparison, a recent study found that during the period of study the COVID-19 mortalilty nationally was 21 percent. ^17^

**Table 4:** COVID-19 Mortality Rates.

| COVID-19 Mortality Rates |  |
| --- | --- |
| Mortality rate in the COVID-19 palliative care population | 33.6% |
| Mortality rate in the non-COVID-19 palliative care population | 28.3% |
| Excess mortality | 5.3% |
| Mortality rate in the COVID-19 non-palliative care population | 9.9% |
| Mortality in the non-COVID-19 non-palliative care population | 5.7% |
| Excess mortality | 4.2% |

In the early days of the pandemic, CareOne isolated all patients transferred from a hospital to a CareOne facility for two weeks to stop the spread of SARS-CoV2. We found that this CareOne containment strategy and the two-week isolation period was effective in reducing transmission hospital acquired infection.

### Causal Factors Reducing Mortality Risk

The causal inference analysis asked counterfactual questions to determine if any treatment provided to COVID-19 patients had a direct causal effect on reducing mortality risk on a case-by-case basis. Such questions posed by the analysis asked whether if patients receiving specific treatments earlier would have had a lower risk of death even if they had not received such care, and conversely, whether people who did not receive treatment would have had a lower risk of death if they had received the treatment.

DAGs were used to eliminate potential confounders such as co-morbidities, severity of illness as measured by oxygen saturation levels, clinical or radiographic presence of pneumonia and multi-organ failure as measured by albumin, alanine transaminase (ALT) and creatine levels.

Figure 1 shows the comprehensive DAG used to assess the causal relationships inferred from our analysis. Prosperdtx developed a complete DAG for every patient in the population to identify a minimal set of covariates (as seen in Chart 1 below) followed by a structural approach to develop a simplified DAG to eliminate confounding, collider bias, selection bias, etc. The simplified DAG of the CareOne patient data set is found in Chart 2 below.

**Figure 1.**
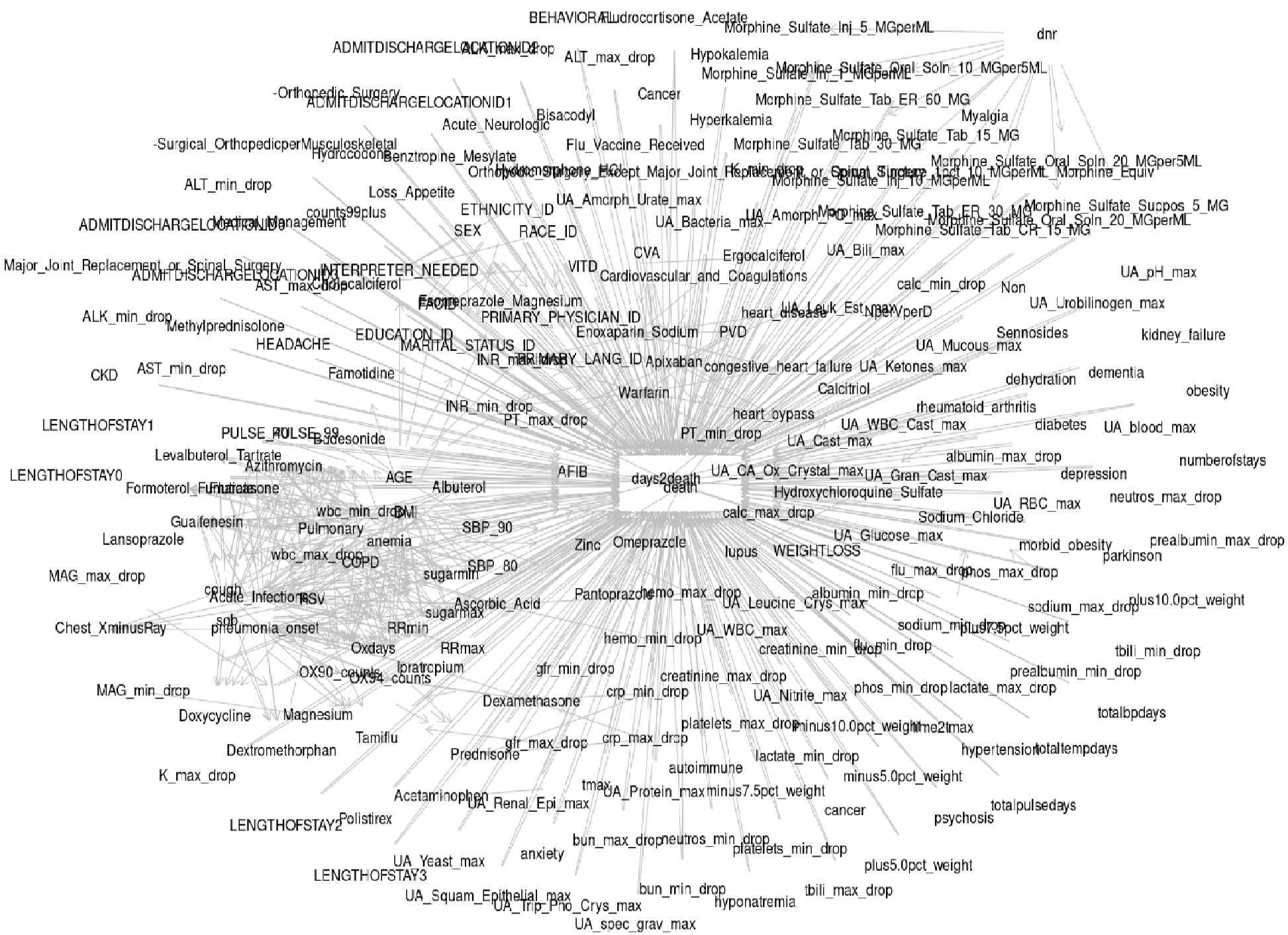
Comprehensive Directed Acyclic Graph (DAG) for Counterfactual Analysis.

**Figure 2.**
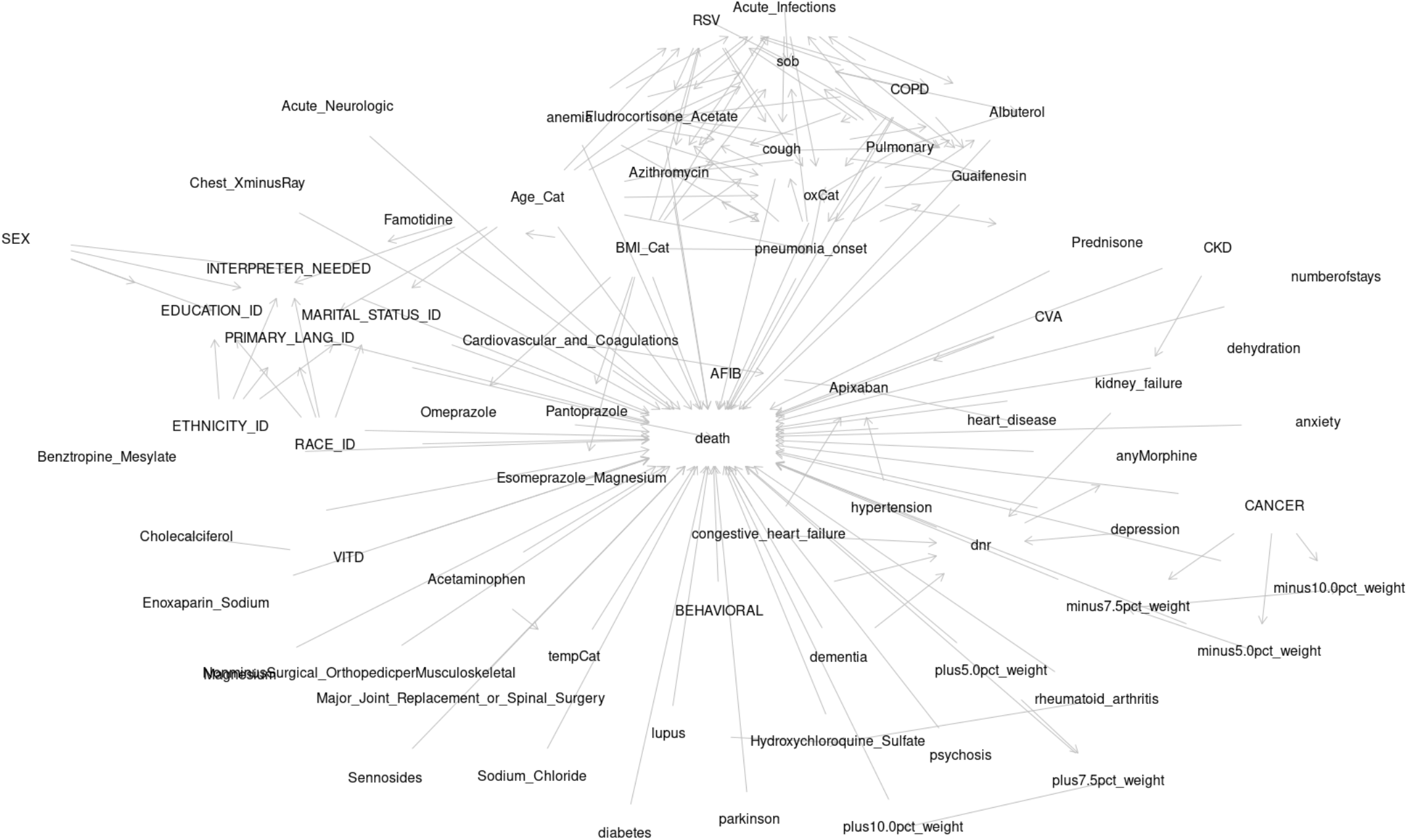
Simplified Directed Acyclic Graph (DAG) for Counterfactual Analysis.

Table 5 displays the results of counterfactual analysis conducted using DAGs to determine what, if any, treatment provided to all COVID-19 positive residents caused a reduction in mortality risk. We also estimated the average treatment effect (ATE) for each intervention. The ATE measures the expected difference in the outcomes if every patient diagnosed with COVID-19 received a treatment to which we posed the counterfactual question: What if every patient diagnosed with COVID-19 had not received the medication? Using DAGs to eliminate confounders allowed us to estimate that the ATE was a function of the observed data distribution.^18^ Counterfactual analysis of the data was conducted to identify interventions with the greatest average treatment effect (ATE) and predict patient level response. We found that the administration of dextromethorphan, prednisone, apixaban, cholecalciferol and omeprazole, at any level of dosing reported in the EHRs reduced the risk of mortality. Table 5 shows that the use of dextromethorphan, prednisone, apixaban, cholecalciferol and omeprazole had average treatment effect estimates across all COVID-19 positive residents of 23.2%, 15.5%, 11.5%, 10.5% and 10.4% and reductions in risk of mortality, respectively. We found that the administration of 800 mg of hydroxychloroquine sulfate (604 unique patients received an administration) did not increase mortality risk as some studies have claimed^19^ nor did it reduce mortality risk.

**Table 5.** Predicted Average Treatment Effect (ATE) on Mortality.

| Medication | % Reduction in mortality risk | Lower confidence interval range 95% | High confidence interval range 95% |
| --- | --- | --- | --- |
| Omeprazole | -0.104 | -0.262 | 0.046 |
| Dextromethorphan | -0.232 | -0.592 | 0.116 |
| Apixaban | -0.115 | -0.278 | 0.062 |
| Cholecalciferol | -0.105 | -0.283 | 0.067 |
| Prednisone | -0.155 | -0.432 | 0.103 |
| Benztropine Mesylate | -0.134 | -0.389 | 0.107 |
| Enoxaparin Sodium | -0.069 | -0.218 | 0.065 |
| Sennosides | -0.063 | -0.303 | 0.187 |
| Guaifenesin | -0.027 | -0.187 | 0.129 |

We conducted an independent component analysis (ICA) of every lab test value to eliminate the possibility of latent variables (Table 6). Using ICA, we were able to determine whether an individual test at a specific measurement influenced the causal model. We ran a 2-way analysis of every lab value against all others and found that no lab value was more predictive of mortality than any other. While some lab values were associated with higher or lower risk than others, they did not influence mortality risk in a causal manner. Further, our component analysis showed that no lab value, independently or in combination, was confounding the impact of specific treatment on mortality rates.

**Table 6:** List of Laboratory Values for Principal Component Analysis.

| Laboratory Values |  |  |
| --- | --- | --- |
| Albumin | Creatinine MAX | Potassium MAX |
| ALK MAX | Creatinine MIN | Potassium MIN |
| ALK MIN | CRP | Platelets MAX |
| ALT MAX | Hemoglobin MAX | Platelets MIN |
| ALT MIN | Lactate | Prealbumin |
| AST MAX | Magnesium MAX | Respiratory rate |
| AST MIN | Magnesium MIN | White blood cells MAX |
| BUN MAX | Max Temperature | White blood cells MIN |
| BUN MIN | Phosphorous |  |

## Discussion

Causal inference analysis showed that specific medications independent of other medications reduced COVID-19 mortality risk among nursing home patients regardless of their comorbidities, demonstrating the utility of observational, longitudinal patient-level data to identify causal relationships between patients, treatments, and outcomes. We can more effectively control for the confounders (a variable, often hidden, that influences the relationship between a dependent variable and independent variable) and collider bias (a third variable is caused by the relationship between an intervention and outcome) using causal inference. Electronic medical records (EHRs) often miss data for one or more variables as was the case in this study. The use of DAGs allowed us to identify when ‘missingness’ might be hiding variables that could bias estimate. However, when we were unable to fully account for missing data in every patient record, for which conditional independence evaluation identified the existence of latent variables, causal relationships were not assessed. However, we showed that causal inference, combined with existing machine leaning methods is an efficient and reliable tool for establishing the causal relationships needed to optimize treatment and personalize medicine. It is possible to do so at scale using automated causal structure learning and to eliminate confounders and selection bias more rapidly and effectively.^20^

Considering the emergence of new COVID-19 variants and the fact that anti-viral medications and vaccines may not be effective in preventing or controlling the disease in all cases, identifying treatment choices that can improve outcomes in all patients, regardless of demography, other diseases, care settings, is critically important. Interventional models derived from causal inference can be used to extrapolate “findings across domains (i.e., settings, populations, environments) that differ both in their distributions and in their inherent causal characteristics.”^21^ Our study also highlights the value of supportive care and its role in achieving optimal health of critically and chronically ill patients. As more care is delivered at home or remotely outside of a healthcare facility, applying causal inference to supportive care in these settings for patients may lead to improved outcomes.

At the same time, we look forward to the development of causal inference methods to generate personalized care pathways that can be dynamically updated to optimize health as patient personal and clinical conditions change. At the outset, we need tools to identify changes in the type, timing, and frequency of care that may have a causal effect on well-being. For example, Marie Davidian pioneered the development of individualized dynamic treatment regimens and created software to allow the use of average treatment effects to estimate optimized treatment outcomes at an individual level.^22^ Finally, there is a need for the deployment of technologies that allow people to create fully integrated health records that can be updated over time. Such data could be collected from electronic medical records, genomic information, wearables, remote monitoring devices and patient surveys. Longitudinal real-time information, combined with causal analysis could be used to make supportive care both prospective and personalized.

## Conclusions

Nursing homes not only dealt with higher rates of Covid-19 infection and death among residents, in several states, patients with COVID-19 from other care settings were taken in by long term care facilities. Diagnosis and treatment decisions were often made without sufficient resources, without an existing body of clinical knowledge for how to best are for a heterogenous group of patients who were also critically ill and without the time and resources to conduct randomized prospective trials to measure treatment effect.

We used causal inference to establish the certain causal and effect relationships of patient treatments and admission policies that providers could use to guide care under such circumstances. This study shows that causal inference can be used to identify important clinical practices for optimizing patient outcomes in various care settings and in people with other conditions. As Pearl has observed, “the entire enterprise known as “personalized medicine” and, more generally, any enterprise requiring inference from populations to individuals, rests on counterfactual analysis and AI now holds the key theoretical tools for operationalizing this analysis.”

Machine learning not only enables the collection and organization of patient level data from many sources, but it can also be used to automate the generation of individual treatment decisions over time as a patient’s condition and circumstances change. When faced with variations of a novel virus, the capacity to quickly determine what supportive care can reduce the risk of death may be as valuable as predicting who is likely to die at a population level.

## Data Availability

All data produced in the present study are available upon reasonable request to the authors

## Acknowledgments

We are thankful for the insightful writings of Dr. Judea Pearl.

## END NOTES

## Footnotes

1 https://data.cms.gov/covid-19/covid-19-nursing-home-data; https://www.kff.org/other/state-indicator/number-of-nursing-facility-residents/?currentTimeframe=0&sortModel=%7B%22colId%22:%22Location%22,%22sort%22:%22asc%22%7D

2 Covid-19 Severity Classifications and Clinical Characteristics 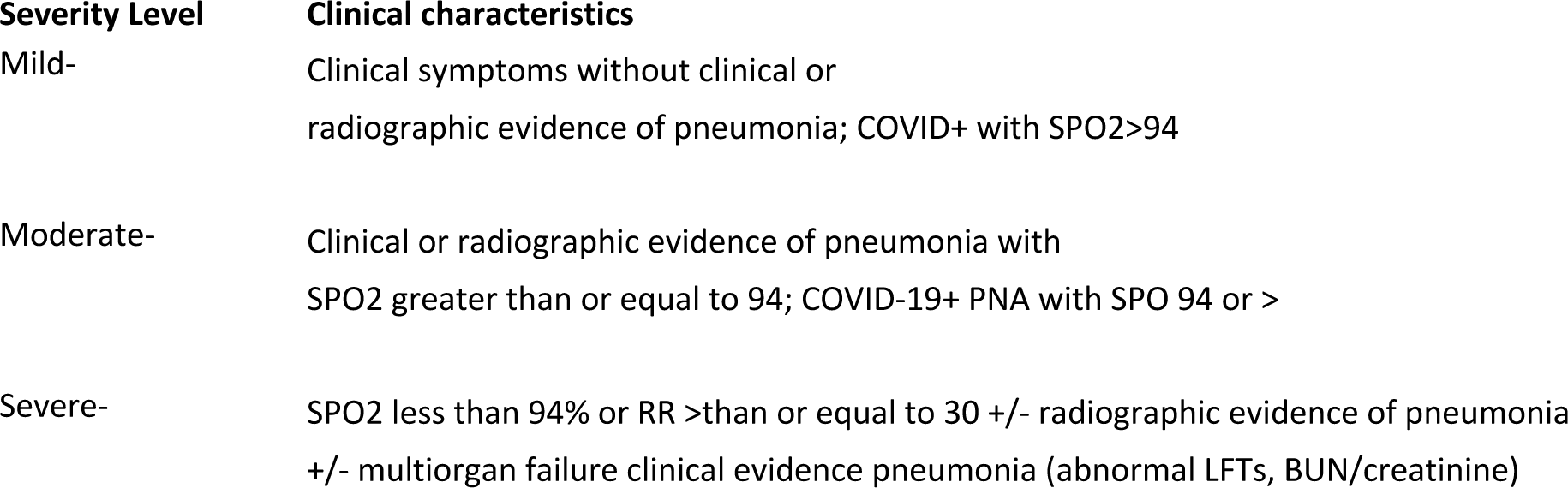

3 Bagchi S, Mak J, Li Q, et al. Rates of COVID-19 Among Residents and Staff Members in Nursing Homes — United States, May 25–November 22, 2020. MMWR Morb Mortal Wkly Rep 2021;70:52–55. DOI: http://dx.doi.org/10.15585/mmwr.mm7002e2external icon.

4 COVID-19 Outbreaks in Long-Term Care Facilities Were Most Severe in the Early Months of the Pandemic, but Data Show Cases and Deaths in Such Facilities May Be On the Rise Again, https://www.kff.org/coronavirus-covid-19/press-release/covid-19-outbreaks-in-long-term-care-facilities-were-most-severe-in-the-early-months-of-the-pandemic-but-data-show-cases-and-deaths-in-such-facilities-may-be-on-the-rise-again/

5 Kosar CM, White EM, Feifer RA, et al. COVID-19 Mortality Rates Among Nursing Home Residents Declined from March to November 2020. *Health Aff (Millwood)*. 2021;40(4):655-663. doi:10.1377/hlthaff.2020.02191

6 Ibid.

7 Pearl, Judea. (2019). The seven tools of causal inference, with reflections on machine learning. Communications of the ACM. 62. 54-60. 10.1145/3241036.

8 Prosperi M, Guo Y, Sperrin M, et al. Causal inference and counterfactual prediction in machine learning for actionable healthcare. Nat Mach Intell. 2020;2(7):369-375. doi:10.1038/s42256-020-0197-y

9 Karthika Mohan & Judea Pearl (2021) Graphical Models for Processing Missing Data, Journal of the American Statistical Association, 116:534, 1023-1037, DOI: 10.1080/01621459.2021.1874961

10 MacKenzie D, **<u>Race</u>**, **<u>COVID Mortality</u>**, **<u>and Simpson’s Paradox</u>** http://causality.cs.ucla.edu/blog/index.php/2020/07/06/race-covid-mortality-and-simpsons-paradox-by-dana-mackenzie/

11 Pearl, J On the Testability of Causal Models with Latent and Instrumental Variables. *UAI’95: Proceedings of the Eleventh conference on Uncertainty in artificial intelligence*

12 Oprescu, M., Syrgkanis, V., & Wu, Z. (2019). Orthogonal Random Forest for Causal Inference. *ICML*.

13 Pedregosa et al. Scikit-learn: Machine Learning in Python, JMLR 2011;12(Oct), pp. 2825-2830.

14 Microsoft Research. EconML: A Python Package for ML-Based Heterogeneous Treatment Effects Estimation. https://github.com/microsoft/EconML, 2019. Version 0.x.

16 ‘The virus isn’t a death sentence’: New Jersey facility takes on COVID-19 with success, https://www.mcknights.com/news/the-virus-isnt-a-death-sentence-new-jersey-facility-takes-on-covid-19-with-success/?utm_source=newsletter&utm_medium=email&utm_campaign=MLT_DailyUpdate_20200506&hmSubId=EEz9pWyN9lk1&hmEmail=O6MNHPLT7RuoX9DsnPHLFQBSfTvWqkiT0&email_hash=dbb217ac7e178edc19e97b3ca1395545&mpweb=1326-9138-204503. Also, https://www.newjerseyhills.com/hanover_eagle/news/update-nursing-home-shut-for-covid-19-patients-moved-to-careone-in-whippany/article_2e01ba5d-e1cb-5010-8c9d-e0c0bb4d6391.html

17 Tarazi WW, Finegold K, Sheingold SH, Wong Samson L, Zuckerman R, Bosworth A. COVID-19-Related Deaths And Excess Deaths Among Medicare Fee-For-Service Beneficiaries. *Health Aff (Millwood)*. 2021;40(6):879-885. doi:10.1377/hlthaff.2020.02521

18 Balzer LB, Petersen ML, van der Laan MJ; SEARCH Collaboration. Targeted estimation and inference for the sample average treatment effect in trials with and without pair-matching. *Stat Med*. 2016;35(21):3717-3732. doi:10.1002/sim.6965

19 Arabi YM, Gordon AC, Derde LPG, et al. Lopinavir-ritonavir and hydroxychloroquine for critically ill patients with COVID-19: REMAP-CAP randomized controlled trial. *Intensive Care Med*. 2021;47(8):867-886. doi:10.1007/s00134-021-06448-5

20 Prosperi M, Guo Y, Sperrin M, et al. Causal inference and counterfactual prediction in machine learning for actionable healthcare. Nat Mach Intell. 2020;2(7):369-375. doi:10.1038/s42256-020-0197-y

21 Bareinboim E, Pearl J. Causal inference and the data-fusion problem. Proc Natl Acad Sci U S A. 2016 Jul 5;113(27):7345-52. doi: 10.1073/pnas.1510507113.

22 Hager R, Tsiatis AA, Davidian M. Optimal two-stage dynamic treatment regimes from a classification perspective with censored survival data. *Biometrics*. 2018;74(4):1180-1192. doi:10.1111/biom.12894

